# Three doses of the BNT162b2 vaccine confer neutralising antibody capacity against the SARS-CoV-2 B.1.1.529 (Omicron) variant of concern

**DOI:** 10.1101/2021.12.23.21268316

**Authors:** Kevin K. Ariën, Leo Heyndrickx, Johan Michiels, Katleen Vereecken, Kurt Van Lent, Sandra Coppens, Pieter Pannus, Geert A. Martens, Marjan Van Esbroeck, Maria E. Goossens, Arnaud Marchant, Koen Bartholomeeusen, Isabelle Desombere

## Abstract

We report the levels of neutralising antibodies against Wuhan, Delta and Omicron variants in healthy individuals pre-infected or not with SARS-CoV-2 and immunized with three doses of the BNT162b2 vaccine. Our observations support the rapid administration of a booster vaccine dose to prevent infection and disease caused by Omicron.

## Introduction

Researchers in Botswana and South Africa identified a new and heavily mutated SARS-CoV-2 variant (B.1.1.529, Omicron) in late November 2021, with 30 amino acid mutations in the Spike protein that are distinct compared to other variants of concern (VOC) Alpha, Beta and Delta [1]. Omicron is characterized by fast spreading in previously vaccinated populations, suggesting Omicron’s ability to evade vaccine-induced immunity [2] and therapeutic monoclonal antibody therapy [3]. Several recent pre-print studies have indeed confirmed a substantial reduction in neutralising antibody activity against Omicron in small-scaled studies including previously infected individuals, fully vaccinated individuals, recipients of third booster doses of BNT162b2 or mRNA-1273 and individuals with hybrid immunity (infection followed by vaccination) [4-9]. The common trend from these first laboratory-based assessments is that the potency to neutralise Omicron is reduced by approximately 40-fold (20-200-fold depending on the study) compared to the original Wuhan D614G virus. Here, we have assessed the levels of neutralising antibodies (nAb) against the original Wuhan strain, Delta (B.1.617.2) and Omicron (B.1.1.529) VOC of 30 sera collected from individuals infected with SARS-CoV-2 prior to vaccination (Group 1), from infection-naïve individuals after three doses of BNT162b2 (Group 2) and from previously-infected individuals after three doses of BNT162b2 (Group 3).

## Methods

### Omicron variant isolation and sequence confirmation

An Omicron virus isolate was made from a nasopharyngeal (NP) swab collected from a patient with sequence-confirmed Omicron infection returning from a stay in the Republic of South Africa. An additional NP swab was collected 48h after RT-qPCR diagnosis and 24h after sequence confirmation of infection with the Omicron variant. The patient was infected after a complete course of vaccination with mRNA-1273 (Moderna) and presented with mild symptoms (cough, sore throat). The NP swap was collected on universal transport medium (UTM) and transferred fresh to the BSL3 laboratory immediately after collection. Two times 200µl of 1/2 diluted sample and 2x 200µl of 1/4 diluted sample (in Vero cell medium with 2% FBS) were added to a 80-100% confluent layer of Vero cells plated 1.5h before in 24-well plates in Vero cell medium with 2% FBS (300.000 cells/well, plates were incubated at 37°C and 7% CO2 before inoculation). Cells were spinoculated for 30 min. at 2500 g and 37°C, then placed again for 10-15 min. in the incubator, after which 800µl of fresh Vero cell medium with 2% FBS was added. Plates were subsequently incubated at 37° and 7% CO2 and cytopathogenic effect (CPE) was scored daily by microscopy. On day 4 post inoculation, CPE was clearly visible in all inoculated wells and virus supernatant from all 4 wells was collected and pooled. Virus was further passaged a second time on Vero cells by adding 2 ml of the freshly collected virus suspension to 95-100% confluently grown Vero cells in a T175 culture flask and incubated for 2h at 37°C and 7% CO2. Next, the cells were washed with PBS, 35ml of fresh Vero cell medium with 2% FBS was added and cells were incubated again at 37° and 7% CO2. On day 4 CPE was clearly visible and virus was collected, centrifuged for 5 min. at 3220 g to remove debris, aliquoted and stored at -80°C. Sequence confirmation regarding Omicron classification was obtained by sequencing the Spike gene on passage 1 of the viral isolate. Viral RNA was extracted using the QIAamp viral RNA mini kit (Qiagen) and the Spike coding sequence was RT-PCR amplified with primer sets COV2-800-39R (CAAAGGCACGCTAGTAGTCGTC), COV2-800-32L (GGGTGTTGCTATGCCTAATCTTTACA), COV2-800-38R (TGCAGTAGCGCGAACAAAATCT) and COV2-800-33L (TGCATGCAAATTACATATTTTGGAGGA), and subsequently sequenced with primers COV2-800-34L (GTTGGATGGAAAGTGAGTTCAGAGT), COV2-800-35L (TTCCGCATCATTTTCCACTTTTAAGT), COV2-800-36L (TGCACAGAAGTCCCTGTTGCTA), COV2-800-37L (TGCAGATGCTGGCTTCATCAAA), COV2-800-38L (CTTCCCTCAGTCAGCACCTCAT), COV2-800-32L (GGGTGTTGCTATGCCTAATCTTTACA), COV2-800-39R (CAAAGGCACGCTAGTAGTCGTC), COV2-800-33L (TGCATGCAAATTACATATTTTGGAGGA), and COV2-800-38R (TGCAGTAGCGCGAACAAAATCT) (adapted from [10]). Using BLAST+, the obtained Spike sequence was found to be identical to a recently published SARS-CoV-2 Omicron sequence from Belgium (GenBank accession number UFO69279.1).

### SARS-CoV-2 whole-virus neutralisation assay

SARS-CoV-2 neutralising antibodies were quantified as previously reported [11,12]. Briefly, serial dilutions of heat-inactivated serum (1/50-1/25600 in EMEM supplemented with 2 mM L-glutamine, 100 U/ml-100 μg/ml of Penicillin–Streptomycin and 2% fetal bovine serum) were incubated during 1 h (37 °C, 7% CO2) with 3xTCID100 of wild type (WT) Wuhan strain (2019-nCoV-Italy-INMI1, reference 008 V-03893), 83DJ-1 (B.1.617.2, Delta) or VLD20211207 (B.1.1.529, Omicron). One hundred µl of sample-virus mixtures and virus/cell controls were added to 100µl of Vero cells (18.000 cells/well) in a 96-well plate and incubated for five days (37 °C, 7% CO2). The CPE caused by viral growth was scored microscopically. The Reed–Muench method was used to calculate the nAb titer that reduced the number of infected wells by 50% (NT50), which was used as a proxy for the nAb concentration in the sample. The Mann-Whitney test was used to compare Wuhan and variant nAb titers.

## Results and discussion

We tested three representative patient groups for neutralising antibody capacity against Wuhan, Delta and Omicron variants of SARS-CoV-2 in a whole-virus neutralisation assay (Figure 1). Group 1 sera (n=10) were obtained from COVID-19 patients hospitalized with severe infection. All patients were infected between 24 Feb 2020 and 27 March 2020 when Wuhan D614G was the only variant circulating and when vaccines were not yet available. Samples tested were collected with a median time after onset of symptoms of 25 days [range 13-46]. Group 2 sera (n=10) were obtained from individuals without a documented previous SARS-CoV-2 infection and 28 days after third dose of BNT162b2. All individuals were vaccinated with a 21-day interval between dose 1 and 2 and received third dose 7 months (median 211 days [207-219]) after dose 2. Group 3 sera (n=10) were obtained from individuals with hybrid immunity, i.e. individuals who have had a previous infection with Wuhan D614G between 24 March 2020 and 11 June 2020, followed by three doses of the BNT162b2 vaccine. Dose 1 and dose 2 were given 21-days apart and the third dose was administered with a median time interval of 8 months (median 261 days [218-290]) after dose 2.

**Figure 1:**
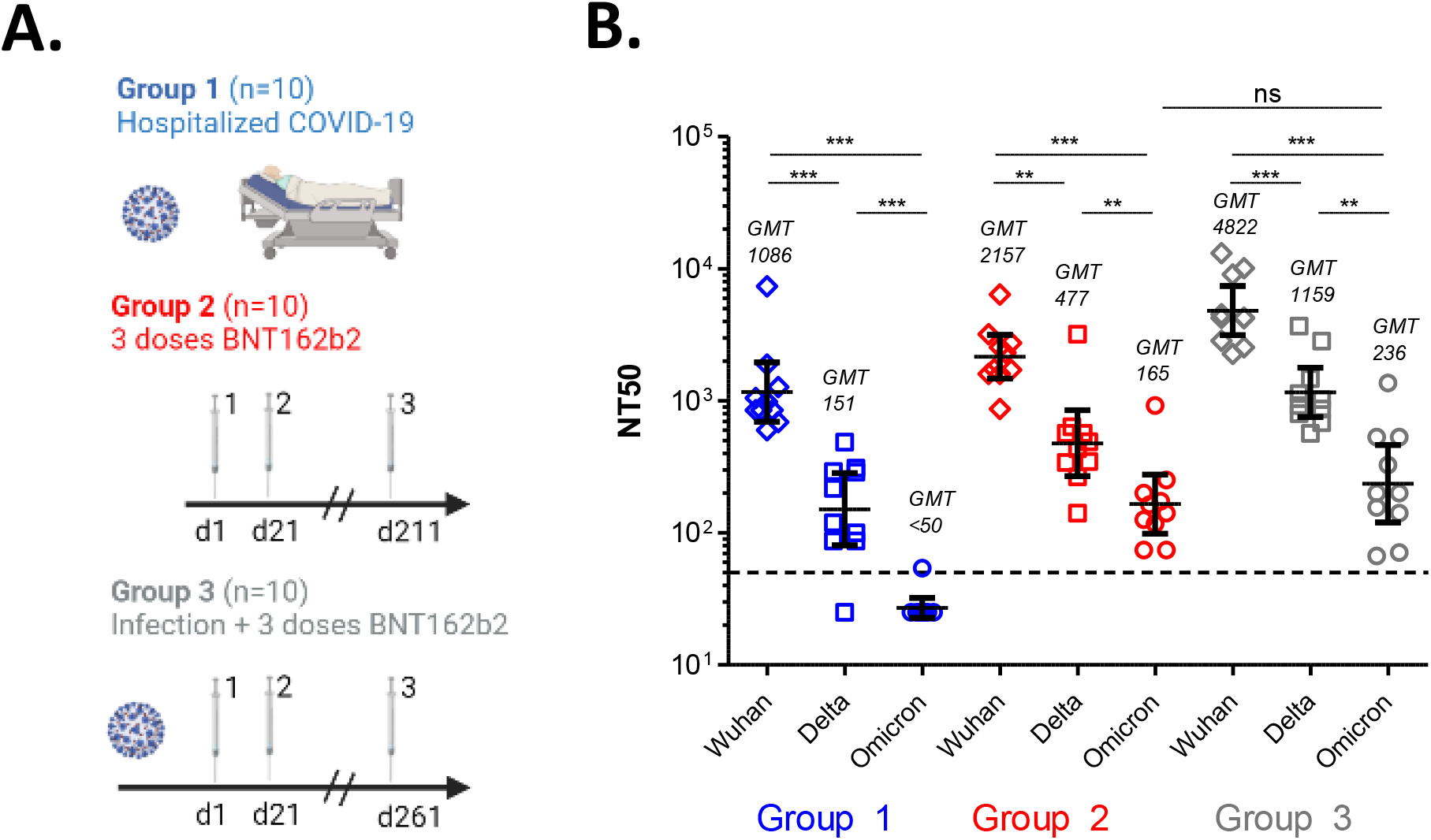
(A) Patient sera characteristics. (B) Neutralising antibody titers (NT50) against Wuhan, Delta and Omicron viruses. GMT (Geometric mean titer), P values Mann-Whitney test: *** (<0.001), ** (<0.01). The dotted line represents the assay cut-off (NT_50_=50).

Individuals with immunity after infection but without vaccination (Group 1) have the lowest nAb responses regardless of the variant tested (Geometric mean titer (GMT)_Wuhan_ 1086, GMT_Delta_ 151, GMT_Omicron_ <50) (Figure 1). Infection-naïve individuals that received three doses of the BNT162b2 vaccine (Group 2) showed significantly higher nAb levels against Wuhan (GMT_Wuhan_ 2157), Delta (GMT_Delta_ 477) and Omicron (GMT_Omicron_ 165). Individuals with hybrid immunity (Group 3) showed the highest NT50 values against Wuhan (GMT_Wuhan_ 4822), Delta (GMT_Delta_ 1159) and Omicron (GMT_Omicron_ 236) variants, but the nAb levels for Omicron were not statistically different between Groups 2 and 3. We observed a significant reduction in NT50 across the three groups for the Delta variant (Group 1: 7.2 fold reduction; Group 2: 4.5 fold reduction; Group 3: 4.2 fold reduction in GMT) and a significantly larger reduction against the Omicron variant (Group 1: >22 fold reduction; Group 2: 13.1 fold reduction; Group 3: 20.5 fold reduction in GMT). This drop in nAb titer is most pronounced for Group 1 (infection/no vaccination), but even for triple vaccinated subjects with (Group 3) or without (Group 2) previous infection, the NT50_Omicron_ is significantly lower than for the Wuhan and Delta strains. Our data confirm the earlier observation that three doses of the mRNA vaccine BNT162b2 results in high neutralising antibody titers for the three variants tested. Although hybrid immunity obtained through infection and subsequent vaccination results in significantly higher nAb levels against Wuhan and Delta, the difference for Omicron is not statistically significant compared to the 3-dose BNT162b2 vaccine schedule.

In this study, we analysed only one critical component of immunity to SARS-CoV-2. Non-neutralizing functions of antibodies and T-cell responses may be less affected by mutations of the Omicron variant and may thereby provide some compensation to immune escape. While multiple studies, including ours, have provided evidence that the Omicron variant can escape naturally acquired and vaccine-induced immune responses, disease severity caused by the variant remains unclear.

In conclusion, our findings confirm that three doses of the BNT162b2 vaccine confers neutralising antibody capacity against Omicron, and that vaccine-induced responses (Groups 2 and 3) outperform naturally-acquired immunity (Group 1). Hybrid immunity (Group 3) results in significantly higher nAb levels against Wuhan and Delta, but not against Omicron. The observation that a 2-dose schedule of BNT162b2 is not sufficient to neutralize Omicron [6,7,9] warrants for rapid administration of a booster vaccine dose to counter infection and limit disease caused by this variant.

## Data Availability

All data produced in the present study are available upon reasonable request to the authors

## Author contributions

KKA wrote the paper. LH, JM, KV, KVL, SC, PP, MVE and KB conducted experimental work and data analysis. KKA, AM, ID conceived the study. KKA and MEG were responsible to obtain the funding. All authors read, edited and approved the final manuscript.

## Acknowledgements

We are grateful to Guido Vanham for critically reviewing the manuscript, to Caroline Rodeghiero and Sarah Houben for logistical and administrative support.

